# The ELEMI healthcare professional Study

**DOI:** 10.1101/2023.03.18.23287312

**Authors:** Gayathri Delanerolle, Yunshan Li, Heitor Cavalini, Katharine Barnard-Kelly, Katheryn Elliot, Vanessa Raymont, Nicola Pluchino, Idaliz Flores, Sohier Elneil, Kingshuk Majumder, Amy Boyd Welsh, Martin Sillem, M. Louise Hull, Rebecca O’Hara, Shanaya Rathod, Ashish Shetty, Jian Qing Shi, Dharani K Hapangama, Peter Phiri

## Abstract

**Introduction:** Endometriosis impacts 1 in 10 women and can be a debilitating disease. Late diagnosis is a major challenge with Endometriosis, contributing further to the exacerbation of symptoms and suboptimal clinical management. Regardless of the commonality of the condition, public awareness and research around endometriosis is severely lacking.

**Methods:** To explore the knowledge base about Endometriosis we developed a digital cross-sectional study. We used the Qualtrics XM platform and developed a questionnaire. The primary objective of the study was to report the understanding of Endometriosis among healthcare professionals in a mental healthcare setting in the UK.

**Results:** We gathered the responses of 144 healthcare professionals, although only 68 participants responded to all questions. Approximately 96% of participants agreed that there is a need for a comprehensive clinical strategy where mental health care services could assist women very early in the pathway. Around 63.1% confirmed awareness of endometriosis although the χ^2^ (p-value=0.158) test showed that the perceived clinical knowledge was not necessarily associated with their profession. Over 92% of participants confirmed that it would be useful to conduct mental health-based research among endometriosis patients.

**Discussion:** It is clear than patients with endometriosis would greatly benefit from a streamlined clinical pathway. It would also have financial benefits to the NHS, including less visits to A&E. The absence of comprehensive knowledge and understanding amongst healthcare professional on endometriosis leads to delayed diagnosis and treatment, exacerbating their psychological and physiological symptoms.

**Conclusions:** Our study shows the importance of funding mental health research to further add to the body of knowledge in order to develop evidence based clinical practices that are equally acceptable to women with endometriosis. This will have benefits for both the patients, healthcare professional, and also the NHS. It will enable for better treatment pathways and symptom management, aiding the development for improved healthcare policies.

## Introduction

Endometriosis is a common disease characterised by the presence of endometrial-like tissue outside the uterus, which leads to adhesions and fibrosis as the tissue breaks down and regenerates in response to cyclical reproductive hormones^1^. Key symptoms among women with endometriosis include chronic pelvic pain, dysmenorrhoea, period-related or cyclical urinary and gastrointestistinal symptoms, and dyspareunia. Many women report depression and anxiety, in addition to a variety of mental health disturbances that may require clinical management^1^. Women with endometriosis are also at high risk of developing other health problems and have an increased life-time risk of health multimorbidity and polypharmacy to manage this condition long-term^2^.

An estimated 1.5 million women are likely to have endometriosis in the United Kingdom (UK), which is similar to the estimates for asthma or diabetes^3^. Endometriosis UK reported 62% of women between the ages of 16 and 24 years in the UK lack awareness of endometriosis, whilst 74% of men do not know about the disease^4^. The National Health Service (NHS) in the UK provides specialist endometriosis centres in a variety of geographical areas, although these are not always universally accessible to patients. As the NHS has primary, secondary and tertiary care settings, a comprehensive data-linkage healthcare record may not be available to streamline the complex clinical management required for endometriosis care^5^. The initial access point for all endometriosis patients is primary care, where General Practitioners (GPs) provide a provisional clinical diagnosis and treatment plan prior to the initiation of any acute or chronic assessments conducted within secondary and tertiary care settings^5^.

Endometriosis patients in the UK frequently access Accident and Emergency (A&E) services to help with emergent symptoms of disease^4^. Endometriosis diagnosis might be difficult since there is no definite diagnostic test and symptoms vary greatly across people. Laparoscopy, which includes inserting a camera via a tiny incision in the abdomen to see the pelvic organs and any apparent endometriotic lesions, is the gold standard for diagnosis^6^. This technique, however, is invasive and not often easily accessible to patients.

Endometriosis treatment choices are determined by the severity of symptoms, the extent of the illness, and the patient’s age and reproductive objectives. Nonsteroidal anti-inflammatory medicines (NSAIDs) are used to treat pain, as are hormonal treatments such as combination oral contraceptives, progestins, gonadotropin-releasing hormone (GnRH) agonists, and danazol^1,7^. In situations when medicinal treatment is inadequate, surgical intervention may be required, which may include the removal of endometriotic lesions and adhesions, as well as hysterectomy and bilateral salpingo-oophorectomy^1,8^.

Despite the fact that endometriosis is common and has a substantial effect on women’s health, there is a dearth of public awareness and research funding for this ailment. To enhance early identification, diagnosis, and treatment of endometriosis, there is a need for improved education and awareness among the general public, healthcare practitioners, and legislators. Additionally, further studies are required to better understand the disease’s underlying causes, create more effective therapies, and eventually improve the life experience for women with endometriosis.

## Methods

We conducted a cross-sectional survey to explore the understanding of Endometriosis among health providers. Our primary aim was to evaluate and report the understanding of Endometriosis among mental healthcare staff in the UK.

### Recruitment

All healthcare providers are eligible to take part in this study through the online survey. Participants will be invited to participate in the study via multiple media sources including intranet, email invites and social media, newsletters and communication campaigns supported by their organisations, deployed online via the NIHR and social media. All participants will be required to complete the survey at a single time point only

### Inclusion Criteria

- ≥18 years
- Any gender
- Healthcare staff working directly with women who have endometriosis
- Healthcare staff working indirectly with women who may have endometriosis (e.g., women’s health services, tertiary care centres such as IAPT)
- Healthcare staff who have access to a smartphone, tablet or computer to be able to complete the survey online.

### Survey details

An online Qualtrics XM platform survey will approximately take 10 -15 minutes to complete following the completion of consent. The survey consists of demographic details, the Pandemic Stress Index, the Flourishing Scale, and the Compassion Fatigue Scale.

### Ethical Approval

This study received HRA REC approval 21/HRA/3500 prior to the study initiation.

### Demographical analysis

A demographic analysis was conducted to analyse the questionnaire data, using descriptive statistics including percentage, mean, median and standard deviation. Population characteristics of gender, profession, healthcare setting and years of service among the study participants were also analysed.

### Non-parametric statistical test

Non-parametric statistical tests were applied to test whether there was significant difference between responses based on gender, profession and healthcare setting. Chi-Square (χ^2^) tests were used to assess differences in responses amongst professions when the answer was nominal. Wilcoxon-Mann-Whitney and Kruskal-Wallis tests were used (the latter was used for the data with more than two groups). The grouping variables included gender, profession and health care setting (primary, secondary or tertiary). Significance was determined by a p value < 0.05. The analysis was undertaken using the SciPy 1.7.1 package with Python 3.9.7.

We merged three healthcare professional groups, academic scientists, radiologists and researchers, together with the original “other” ((“other” is a group among several professions and is derived from the original “academic scientists”, “radiologists”, “researchers” and “other”.) as one group (still denoted as ‘other’) since the number under each of the groups was too small. We then produced a contingency table to conduct a χ^2^ test to report the difference in opinions between this group and the other groups.

We also ran a post-hoc study to investigate the disparities in endometriosis knowledge across mental healthcare employees and other healthcare experts. Since the number of participants in each group was limited, we combined three healthcare professional groups (academic scientists, radiologists, and researchers) with the original “other” group to form one group (denoted as “other”).

We pooled the different healthcare groups to produce a contingency table to conduct a χ^2^ test to report the difference in opinions between this group and the other groups. The threshold of significance was fixed at p=<0.05. This study was also carried out using Python 3.9.7 and the SciPy 1.7.1 module.

All participants provided informed permission before being permitted to view the survey. The survey was entirely optional, and participants were free to leave at any moment. The University of Bristol Ethics Committee authorised the project.

The questionnaire (See Supplementary document A) was created using current research, and assistance from professionals with endometriosis treatment expertise. It included 26 questions on basic endometriosis knowledge, diagnosis, treatment choices, and the effect of endometriosis on patients’ mental health. Additionally, two validated questionnaires were also included. The first was the ‘Flourishing Scale’, an 8-item summary measure of self-perceived purpose, optimism, and self-esteem. The second was the ‘Compassion Fatigue Scale’, a 13-item scale measuring the psychological, physical, and emotional impact of working within a care role.

The poll was sent out through multiple methods, including social media, email, and professional networks, with a focus on mental healthcare workers in the United Kingdom. From May to July 2021, data was collected, and 727 replies were obtained.

The data was analysed using the above-mentioned statistical software programs to find any significant variations in knowledge and awareness of endometriosis across UK healthcare workers, with a special emphasis on mental healthcare employees.

## Results

The sample included 144 healthcare professionals, however only 68 answered all the questions. Therefore, the rate of missing data for questions ranged from 35.4% to 77.8%.

### Demographical analysis

The demographic analysis of the survey results revealed that the majority of the participants (85.6%) were female, with the nursing profession having the largest representation (45.8%), followed by psychology/psychotherapy (18.3%), and medicine (11.6%). The majority of participants (52.7%) were from secondary care settings, followed by primary care (29.5%) and tertiary care (17.8%). The participants’ average number of years of service was 12.5 years.

### Gender

Of the 144 participants, 78 participants (54.2%) were females, 13 (9.0%) were males, 2 (1.4%) preferred not to say and for 51 participants’ (35.4%) gender information was missing (Figure 1 and Supplementary Table 1).

**Figure 1.**
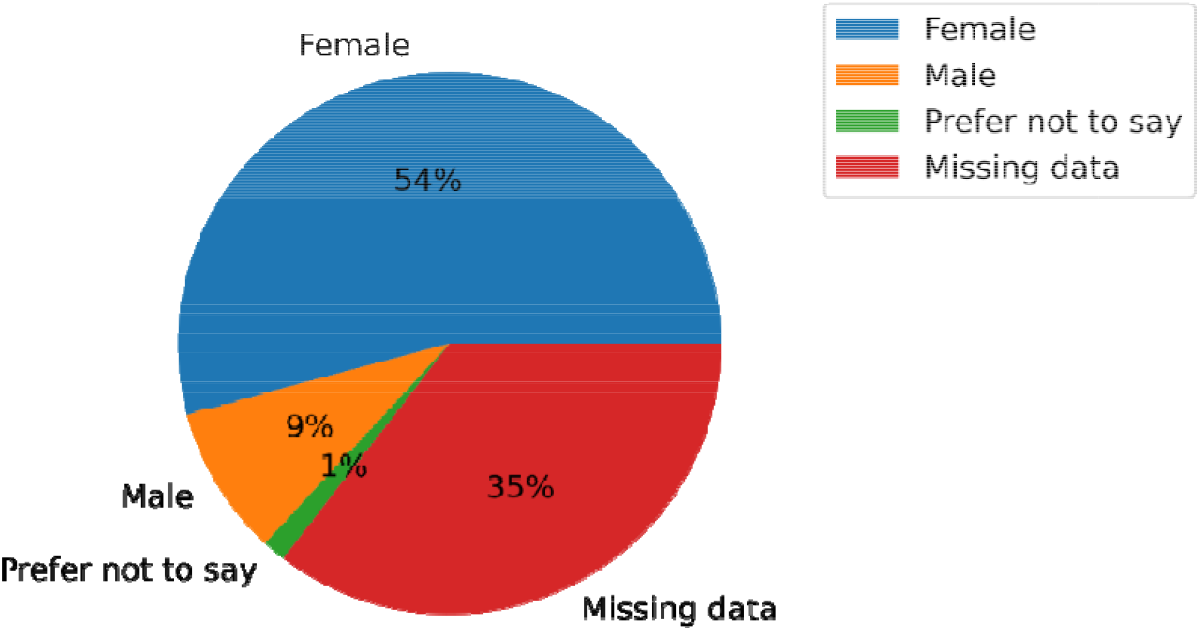
Gender distribution of participants

### Profession

The participants comprised psychiatrists or mental health staff 35 (24.3%), nurses 24 (16.7%), researchers 3 (2.1%),, a radiologist1 (0.7%) an academic scientist 1 (0.7%) and other clinical groups 29 (20.1%) (Figure 2 and Supplementary Table 2). A total of 51 participants’ (35.4%) did not complete the professional information section.

**Figure 2.**
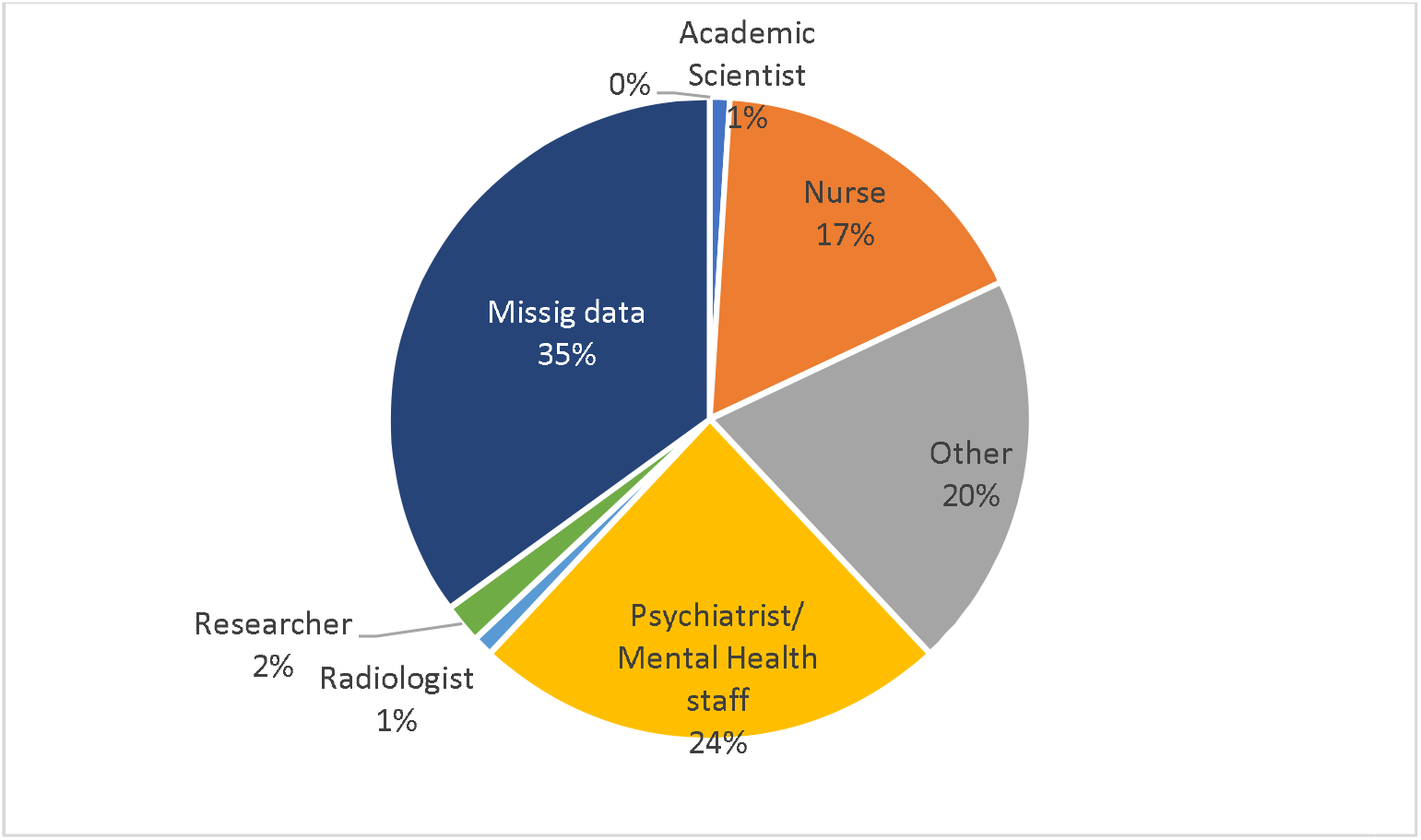
The profession distribution of participants

### Professional experience

Participants had a mean of 102.9 months (SD: 118.8), in their chosen profession (Figure 3). A total of 39 participants’ (27.1%) were working within primary care, 36 (25.0%) at secondary level, 7 (4.9%) were at tertiary level and 11 (7.6%) were at other levels. Fifty one participants (35.4%) did not report this information (Figure 4 and Supplementary Table 3).

**Figure 3.**
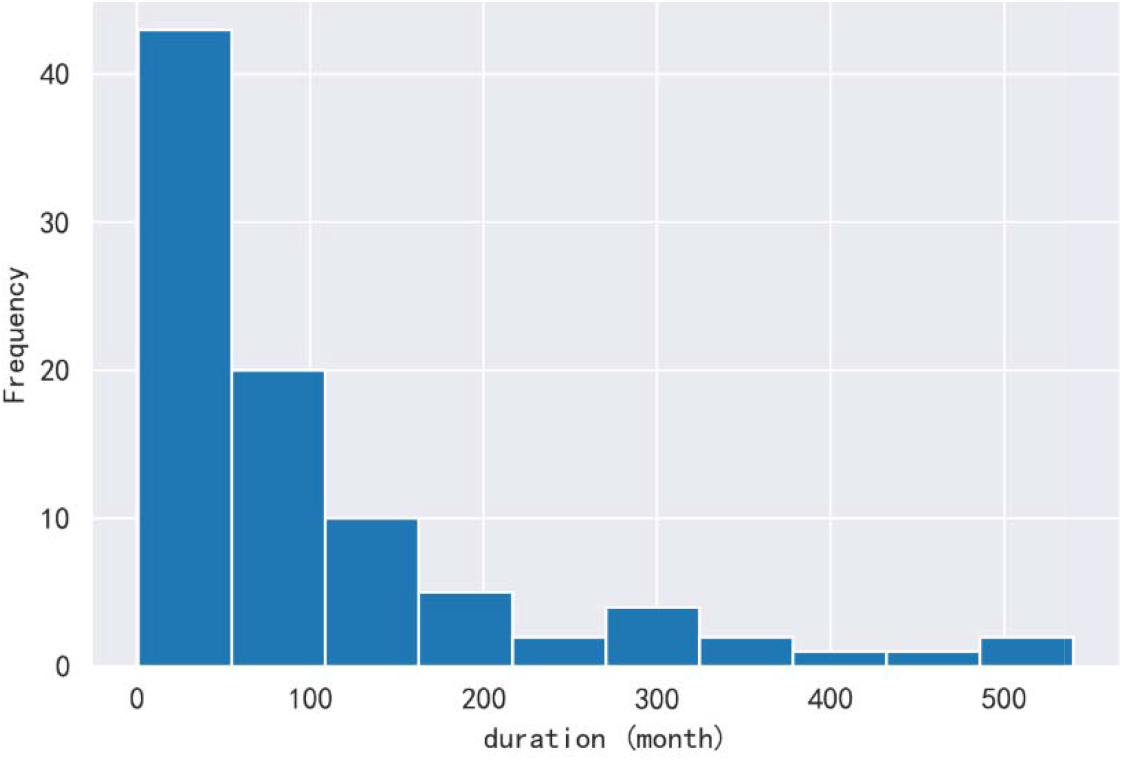
Participant’s duration in chosen profession

**Figure 4.**
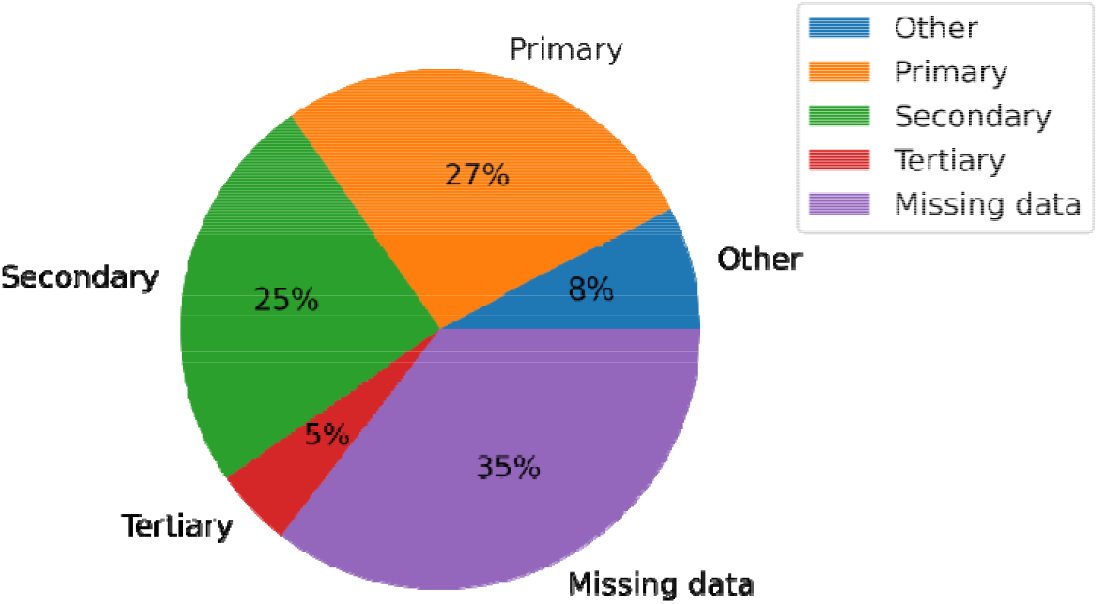
Healthcare setting of participants

### Non-parametric statistical tests

Three primary themes were reported by all the participants in relation to Endometriosis associated knowledge and practice.

### Perceived Clinical Knowledge

Approximately 63.1% of the participants confirmed that they knew about Endometriosis and were confident in their level of clinical knowledge (Supplementary Table 4 and Figure 5). The test showed that perceived clinical knowledge was not significantly linked to their profession (p-value=0.158).

**Figure 5.**
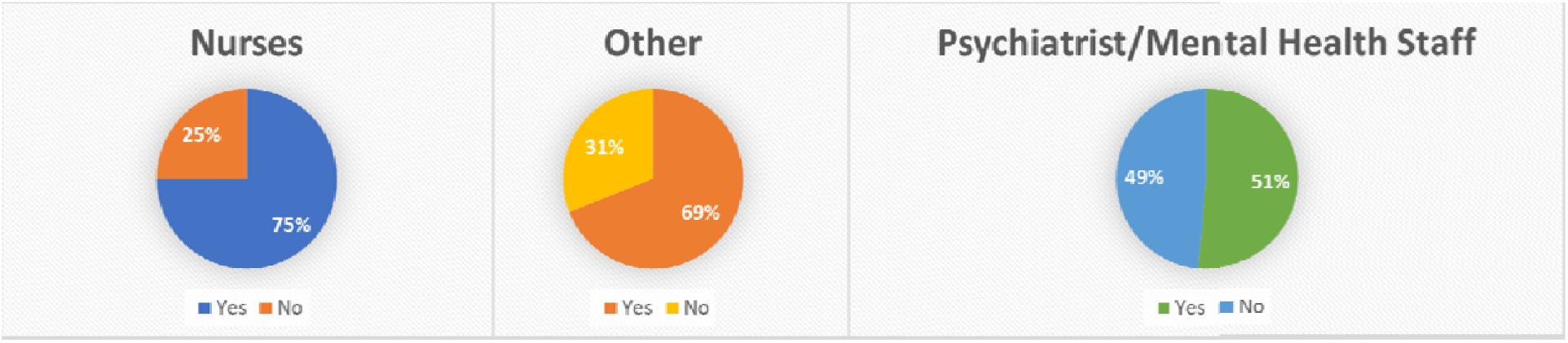
Contingency chart indicating agreement with the questions: “Do you know what Endometriosis is? Are you comfortable with your level of clinical knowledge around it?”

### Endometriosis Clinical Pathway

A total of 96% of the participants agreed that they feel Endometriosis patients would benefit from a comprehensive clinical pathway that could be implemented across primary, secondary and tertiary care (Figure 6), whilst 4.0% did not agree with this view. The test of p-value of 1.0 in Supplementary Table 5 shows the opinions in different groups are consistent.

**Figure 6.**
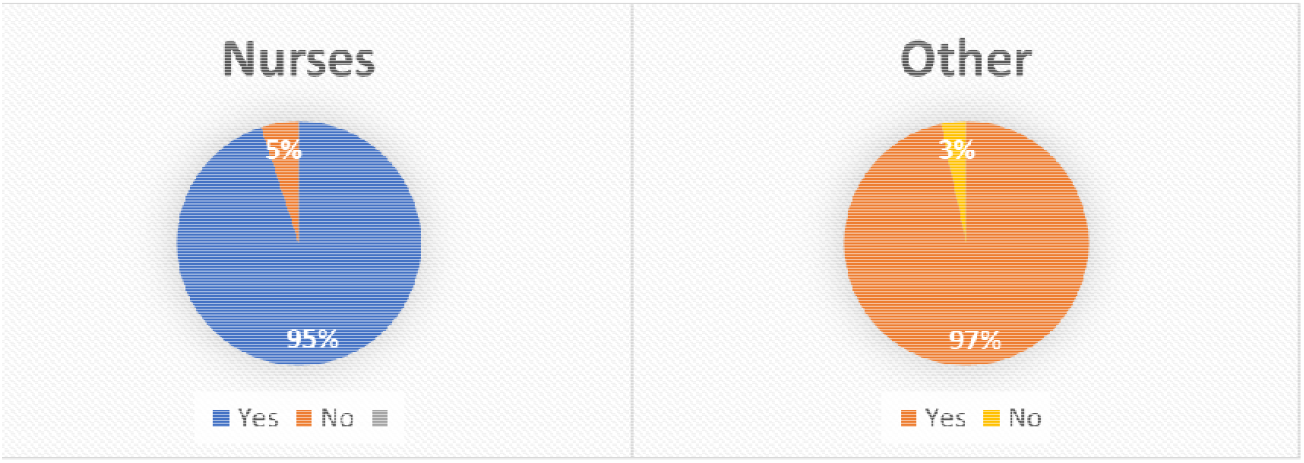
Contingency graph indicating agreement with “Do you think patients with endometriosis would benefit from more comprehensive pathway that could be universally implemented across primary, secondary and tertiary care?”

### Mental Health Research

Approximately 92.3% of all participants agreed that it would be useful to conduct mental health-based research among women with endometriosis to better understand, navigate and develop clinical, and non-clinical interventions; whilst 7.7% d d not agree with this statement (Figure 7 and Supplementary Table 6). The test showed a p-value of 0.556 and that the views may not necessarily be linked to their profession.

**Figure 7.**
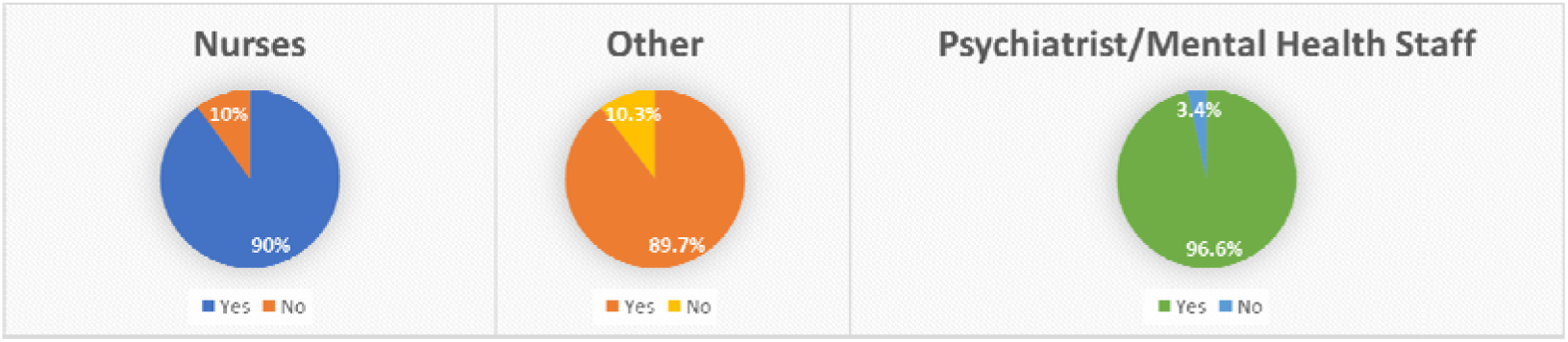
Contingency chart indicating agreement with “Do you think it would be useful to conduct mental health-based research for endometriosis women so that better clinical and/or non-clinical interventions can be developed?”

### Satisfaction

Participants reported varying levels of satisfaction with the support provided by local services to endometriosis patients. A total of 10.4% of participants were very dissatisfied, 12.5% dissatisfied, 54.2% remained neutral, 20.8% satisfied and 2.1% very satisfied Although there were differing views among the different categories of professionals (Table 1) this did not reach statistical significance (p-value = 0.158).

**Table 1.** Satisfaction based on the services provided to endometriosis patients by profession

| Profession | Very dissatisfied |  | Dissatisfied |  | Neutral /Not sure |  | Satisfied |  | Very satisfied |  | Total |
| --- | --- | --- | --- | --- | --- | --- | --- | --- | --- | --- | --- |
| Nurse | 4 | 20.0% | 3 | 15.0% | 9 | 45.0% | 4 | 20.0% | 0 | 0.0% | 20 |
| Psychiatrist /Mental health staff | 1 | 3.6% | 3 | 10.7% | 17 | 60.7% | 6 | 21.4% | 1 | 3.6% | 28 |
| Total | 5 | 10.4% | 6 | 12.5% | 26 | 54.2% | 10 | 20.8% | 1 | 2.1% | 48 |

Note: the p-value of test was 0.158, showing no significant difference of satisfaction between different professions; the number of missing answers was 95 leading to a missing rate of 66.0%.

### Participant mental health and wellbeing

There were two scale questionnaires (Flourishing Scale and Compassion Fatigue Scale) to measure participants’ mental health which included their quality of life. The mean quality of life score for all participants was 45.26 (SD: 7.86) out of 56 (Figure 8) and the higher the score was the better life quality it indicated. To determine whether there was significant difference of the scale scores between different genders, professionals and setting, Wilcoxon-Mann-Whitney tests were performed. Ther was a significant difference in quality-of-life scores between different professions (p-value: 0.001, Figure 9). The means score among the nursing staff was 48.59, psychiatrist/mental health staff 47.11 and researchers 47; which was higher than that of the radiologist 32. There was no statistically significant difference based on gender (p=0.231) or the setting (p=0.429, Figure 9).

**Figure 8.**
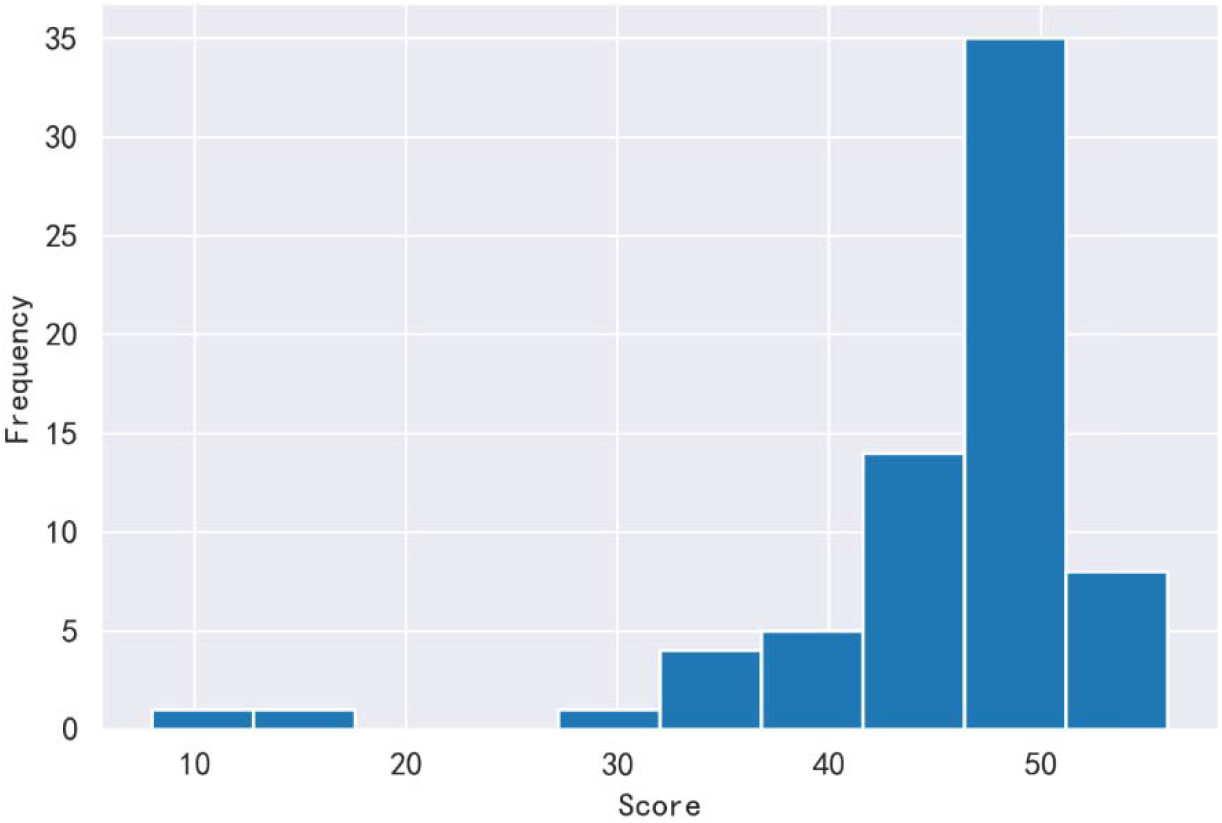
Quality of life scores (there were 75 missing scores leading to a missing rate of 52.1%)

**Figure 9.**
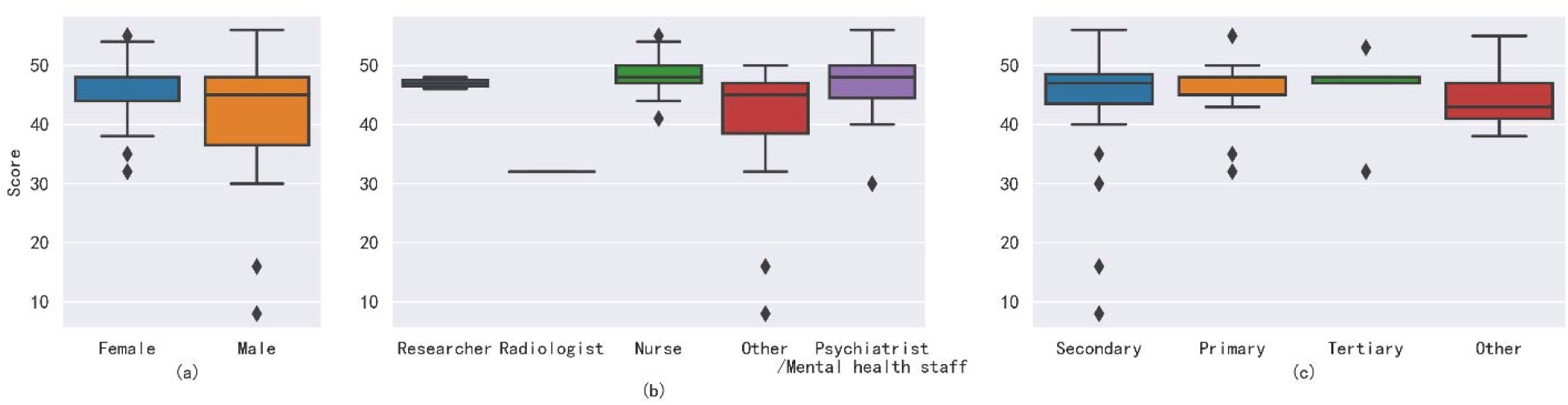
(a) Boxplot illustrating quality of life by gender (those responding “prefer not to say” for gender were not included in analysis). The p-value was 0.231, showing no significant difference between genders. (b) Boxplot of quality-of-life scores by profession. The p-value was 0.001, showing significant difference between professions. (c) Boxplot of quality-of-life scores by health setting. The p-value was 0.429, showing **no** significant difference between levels of local healthcare system.

A mean difficulty of life situation score of 47.38 (SD=19.47) out of 130 was recorded (Figure 10) and the higher the score was the more difficult a life situation it indicated. To determine whether there was significant difference of the scale scores between different genders, professions and settings, a Wilcoxon-Mann-Whitney test or a Kruskal-Wallis test was performed. A p-value of 0.022 was recorded when the scores from different professional groups were analysed, indicating a significant difference in scores between the groups. The mean score among psychiatrists/mental health staff was 50.18 compared to researchers who had the lowest score at 37.5 (Figure 11). There were no significant differences between genders (p-value=0.315) or setting (p-value=0.081, Figure 11).

**Figure 10.**
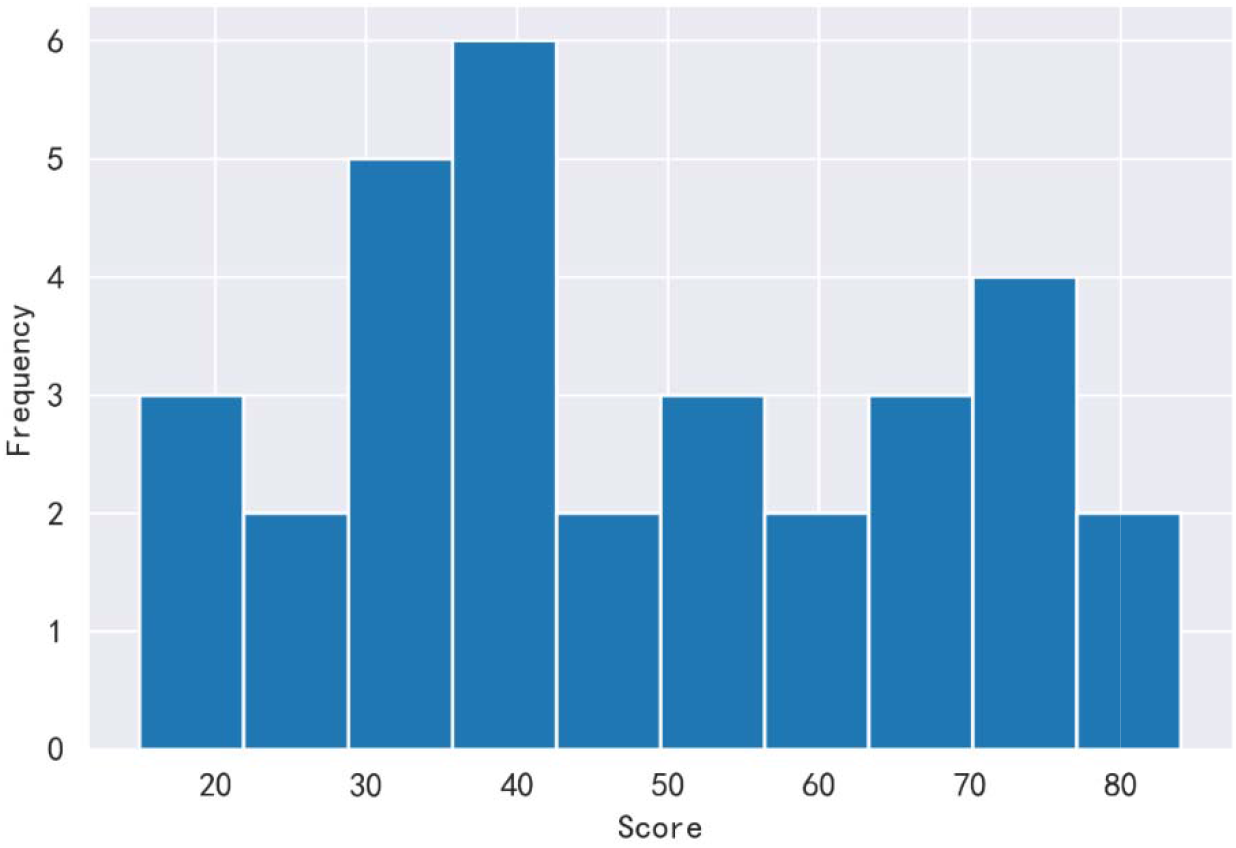
Histogram of scale scores of difficulty of life situation (there were 112 missing scores leading to a missing rate of 77.8%)

**Figure 11.**
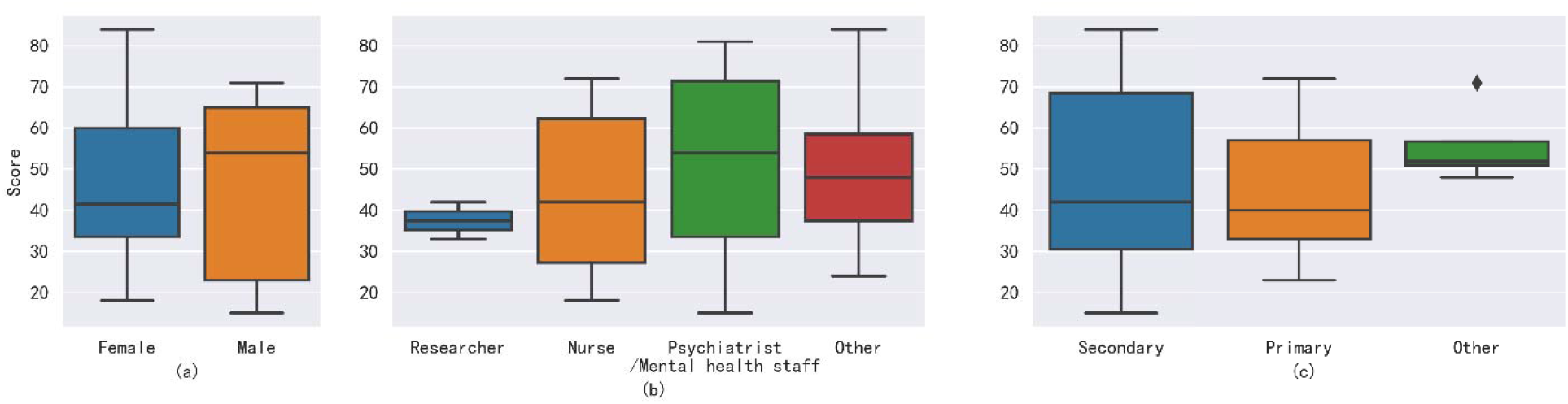
(a) Boxplot of difficulty of life situation by gender (those responding “prefer not to say” for gender were not included in analysis). The p-value was 0.315, showing **no** significant difference between genders. (b) Boxplot of difficulty of life situation by profession. The p-value was 0.022, showing significant difference between professions. (c) Boxplot of difficulty of life situation by setting. The p-value was 0.081, showing no significant difference between levels of local healthcare system.

The study found that there was a general lack of knowledge and a areness of endometriosis among UK healthcare professionals, especially those in mental health. The survey discovered that only 33.8% of individuals had a solid grasp of endometriosis.

- Fewer than half of the individuals correctly identified the most prevalent endometriosis symptoms, which included persistent pelvic pain (47.3%), dysmenorrhea (40.3%), and painful intercourse (36.3%).
- Just 29.5% of those polled were aware that there is no cure for endometriosis, and fewer than half were aware of the different treatment choices.
- As compared to primary care, healthcare personnel in intermediate and tertiary care settings had a greater knowledge of endometriosis.
- There were substantial gender and profession variations in endometriosis comprehension and awareness, with female participants and those in nursing or psychology/psychotherapy having superior knowledge than male participants and those in other professions.
- Respondents perceived that endometriosis had a limited influence on mental health, with just 28.8% of mental health providers noting the increased risk of despair and anxiety in women with endometriosis.

Overall, the research recommends that greater information and training for healthcare professionals, especially those in primary care and mental health, is needed to enhance the treatment and care of endometriosis patients in the United Kingdom.

## Discussion

A key finding of this study is that majority of healthcare providers felt that Endometriosis patients would benefit from a streamlined clinical pathway. This could encompass timely recognition and diagnosis followed with medical treatment (e.g., surgery, medication). Psychological support could also be integrated with this to help with the emotional strain. Manifestations of physical and psychological symptoms can often be managed optimally if the relevant professional staffing groups are aware of the clinical background and ongoing care requirements of individual patients. Better management of patients could aid the NHS from a cost’s perspective as the potential number of A&E visits by Endometriosis patients could be reduced. In addition, having a more streamlined clinical pathway could improve both communication and engagement levels between healthcare professionals and patients.

Endometriosis is recognised by caregivers as a complex disease to manage. More women than men participated in this study. It is important to improve gender representation in research but far more vital to understand if male healthcare professionals are aware of and understand endometriosis. The mental health impact of endometriosis among women indicates the need for better understanding and communication methods, as shown by the ‘Endometriosis in the UK: time for change’ reported published in 2020 by the All-Party Parliamentary Group. Patient advocacy groups have also indicated the need for better engagement and communication with endometriosis patients, especially in relation to their mental health and wellbeing given the potential exacerbation of the condition due to stress.

Patients with endometriosis engage with multidisciplinary teams at primary, secondary and tertiary care settings. The professional background is an important facet to consider exploring potential pathways and training requirements that could be designed in the future. Each of the settings have a variety of localised pathways and access to seek mental health services. The study findings revealed that there were considerable disparities in knowledge and awareness of endometriosis across UK healthcare professionals. Healthcare personnel in secondary and tertiary care settings, for example, have more knowledge and awareness of endometriosis than those in primary care. There were also substantial disparities in endometriosis comprehension and awareness depending on gender and career.

The most common route used in the UK is via GPs based in a primary care setting, although National Institute for Health and Care Excellence does not recommend the use of anti-depressants as a first line treatment for women with endometriosis reporting anxiety and/or depression at present. Given the current clinical pressures within primary healthcare services, often GPs refer patients to Improving Access to Psychological Therapies (IAPT) services for psychological interventions as recommended by the National Institute for Clinical Excellence (NICE) guidelines. Cognitive behavioural therapy (CBT) can often be a first step to support patients who may want to use a non-pharmacological route to secure therapeutic benefit. CBT could also be a sustainable tool that could be used by patients in a manner that is suited to their needs, empowering them to also have improved health seeking behaviours. This may not be a consideration either for the relevant mental healthcare professionals seeing the patients as it is not within their standard health questionnaire obtained at the first visit. Subsequent visits may not have such questionnaires which could prevent a diagnosis of endometriosis being recorded for those women who receive a late diagnosis, further complicating the ability to provide a more bespoke CBT approach. Furthermore, at present mental health care services currently lack endometriosis specific psychological protocols and the complexities with any ongoing acute care may not be taken into consideration when generic approaches are administered to patients. Thereby, the therapeutic benefit may become suboptimal quite quickly.

Another important consideration from this survey is that 92.3% of participants considered that mental health research conducted among endometriosis patients could aid the understanding, management and development of more suitable interventions that can be sustainably used. Often endometriosis patients are in significant pain and could be using analgesics that could negatively influence their overall medium to long-term wellbeing. Developing interventions that are of therapeutic benefit, minimally or non-invasive, and personalised to patient’s requirements could also increase the overall healthcare outcomes in a positive manner among endometriosis patients. To achieve this, extensive clinical research would be required involving key stakeholders (e.g., patients and healthcare providers). This is an important point for funders as there is currently a paucity of funding available for endometriosis and mental health research, yet this seems to be a huge clinical need.

Endometriosis is largely a medical ailment, but it is also connected with a high incidence of anxiety, depression, and other mental health issues. This has the potential to significantly affect patients’ quality of life and overall health outcomes. Healthcare practitioners must be aware of this and adopt a comprehensive approach to endometriosis treatment that considers both the physical and psychological elements of the ailment. Professional inexperience coupled with the awareness of endometriosis was another vital facet the study explored to better understand potential gaps. According to the findings of this survey, 63.9% of participants were unfamiliar with endometriosis, and only 22.8% thought they had adequate expertise to handle individuals with the illness. The research also emphasized the need of training and raising awareness among healthcare workers. To understand and accept the complex needs of endometriosis patients, a cultural transformation may be required as patients have reported concerns where they have felt to have not been heard. Endometriosis is still not commonly taught in medical colleges; therefore some doctors may be unaware of the illness. Participants noted a need for improved endometriosis education and training, as well as the need of having clear clinical standards for its care. This lack of knowledge may lead to delays in diagnosis and effective treatment, which can have a substantial impact on patients’ physical and emotional health. This underlines the critical need for endometriosis education, include increased awareness around endometriosis symptoms, risk factors, diagnosis, and care, as well as impacts on mental health and well-being.

Additionally, the research discovered that healthcare providers may not always consider the effect of endometriosis on fertility and reproductive health. Endometriosis is a prevalent cause of infertility, and people with the illness may need fertility therapy from a professional in order to conceive. Nevertheless, the survey indicated that only 32.8% of healthcare providers were competent in handling endometriosis’s reproductive components, and 43.6% were not confident in addressing fertility difficulties with patients.

## Limitations

One methodological limitation was that the sample size was of a medium scale. Future research would benefit from having a larger sample size to draw more comprehensive conclusions from some χ^2^ tests. Due to this study taking place during the COVID-19 pandemic, there were many challenges due to time constraints and availability of healthcare professionals in the UK.

## Conclusion

This research emphasizes the need of healthcare workers recognizing endometriosis, its influence on patients’ mental health and wellbeing, and its consequences for conception and reproductive health. It is vital that healthcare providers obtain the appropriate information and training in order to properly manage endometriosis patients and give the support and care they need to enhance their overall quality of life. There is a need for a more simplified clinical route for endometriosis patients, which might increase communication and engagement levels between healthcare personnel and patients. Comprehensive endometriosis education and training is required as well as the creation of endometriosis-specific psychological procedures. Healthcare services in the UK would benefit from conducting mental health research among endometriosis women to better understand the complex needs of the patients, alignment of these to develop patient centric healthcare services, improve access to mental healthcare services in areas where services currently exist and sustain these services to a growing population. Improved interim healthcare policies and clinical guidelines should be in place that are more patient centric to meet the demands of the endometriosis patients.

## Data Availability

All data produced in the present work are contained in the manuscript

## Acknowledgements

The authors acknowledge support from Southern Health NHS Foundation Trust.

